# No evidence of an association between graft-versus-host disease and BK polyomavirus viral proteins LTAg and VP1– a pilot investigation

**DOI:** 10.64898/2026.11.17.26368216

**Authors:** Shabnam Asadikomeleh, Thomas Neumann, William Krüger, Silvia Ribback, Laila Schneidewind

## Abstract

**Introduction:** This is an immunohistochemical (IHC) pilot study of adult patients who underwent allogeneic stem cell transplantation (alloSCT) for hematological disease and had BKPyV reactivation (BKPyV viruria) as well as graft-versus-host-disease (GvHD) to investigate, if there is an association between these two diseases.

**Methods:** The study was prospectively registered at the German Clinical Trials Register (https://drks.de/search/en/trial/DRKS00039127/details). The full protocol is available with this link.

**Results:** Seven (58.3%) of the identified patients were treated in our prospective clinical trial regarding urological complications in alloSCT. All of them had BKPyV viruria during inpatient treatment for the transplantation and developed acute GvHD of the skin. Five (41.7%) of the patients were recruited in our prospective study of long-term survivors (more than five years) of alloSCT. All of them had BKPyV viruria as well as chronic GvHD of the skin at time of their examination. Acute myeloid leukemia was the most frequent underlying indication for alloSCT (n=7; 58.3%), followed by Non-Hodgkin-Lymphoma (NHL; n=2; 16,7%). Interestingly, IHC was negative for LTAg as well as VP1 in all patients. Therefore, it must also be stated that both virus proteins were negative in IHC for acute as well as chronic GvHD of the skin. Consequently, no further molecular investigations were realized.

**Conclusion:** There is no evidence for an association between the viral proteins LTAg and VP1 of BKPyV and skin GvHD in immunohistochemistry. Due to the pilot character of our investigation, the results should be interpreted with great caution.

## Introduction

Since several decades it has been controversially discussed, if graft-versus-host-disease (GvHD) is a risk factor for BK polyomavirus (BKPyV) associated hemorrhagic cystitis (HC) in adult allogeneic stem cell transplantation (alloSCT) [1; 2; 3; 4; 5; 6; 7; 8; 9; 10; 11; 12]. In a recent comprehensive meta-analysis, acute GvHD (OR 2.73; 95% CI 2.02 – 3.69; p<0.0001; I^2^=46%) as well as chronic GvHD (OR 1.71; 95% CI 1.12 – 2.60; p=0.01; I^2^=0%) have been identified as risk factors for BKPyV-HC in that case [13]. However, it has also been suggested that BKPyV-HC could also be a generalized epithelial disease due to reactivation of the virus [14]. Moreover, GvHD generally involves multiple systems and can also involve the urothelium of the bladder, where BKPyV usually persists [13; 15; 16; 17; 18; 19; 20]. Therefore, the question arises: Is there a causal relationship between the infection/reactivation with BKPyV and GvHD, or are the reasons for the described as-sociation are primarily of immunological nature [13]. In detail, is BKPyV mostly replicating due to extensive immunosuppressive therapy against GvHD or is the virus also replicating in GvHD tissue and therefore involved in pathogenesis or progression of this disease? Consequently, we conducted an immunohistochemical (IHC) pilot study of adult patients who underwent alloSCT for hematological disease and had BKPyV reactivation (BKPyV viruria) as well as GvHD. Initially, also further molecular analysis was planned.

## Methods

From our previously conducted and published prospective clinical studies on urological complications in alloSCT [21; 22], we were able to identify 12 patients who had both BKPyV viruria and GvHD of the skin, and for whom paraffin blocks from the histolog-ically-proven GvHD were available for further IHC analysis. Before starting the study, we obtained the approval of the local ethics review board at the University Medical Center Greifswald (BB 264/25 from December 21, 2025) and the study was prospectively registered at the German Clinical Trials Register (https://drks.de/search/en/trial/DRKS00039127/details). All our studies were designed according to the guidelines in the synthesis of qualitative research (ENTREQ) found on the equator network.org, an international initiative providing robust reporting guide-lines [23]. The full study protocol is available via the link and DRKS ID. IHC was per-formed of 5 μm sections for the early large tumor antigen of BKPyV (LTAg, anti-SV40 LTAg, Santa Cruz Biotechnology, Dallas, Texas, USA, sc-53448, mouse monoclonal) and of the late capsid protein (VP1, anti-SV40-VP1, Abcam, Cambridge, Great Britain, ab-53977, rabbit polyclonal) according to our published protocol [24]. Probes from bi-opsy proven BKPyV-nephropathy were used as positive controls and slides from our infection experiments [24] as well as skin probes from patients of our studies with BKPyV viruria, but without GvHD, served as negative controls. Then IHC was evaluated with standardized settings with a DMI 6000B microscope (Leica, Wetzlar, Germany) and the Microsoft Image Composite Editor program.

## Results

Seven (58.3%) of the identified patients were treated in our prospective clinical trial regarding urological complications in alloSCT [21]. All of them had BKPyV viruria during inpatient treatment for the transplantation and developed acute GvHD of the skin. Five (41.7%) of the patients were recruited in our prospective study of long-term survivors (more than five years) of alloSCT [22]. All of them had BKPyV viruria as well as chronic GvHD of the skin at time of their examination.

For the whole study population of this investigation: Acute myeloid leukemia was the most frequent underlying indication for alloSCT (n=7; 58.3%), followed by Non-Hodgkin-Lymphoma (NHL; n=2; 16,7%). Furthermore, eight (66.7%) patients were male and four (33.3%) were female.

Regarding the seven patients of our prospective clinical trial about urological complications in alloSCT, NHL was the most frequent underlying disease (n=2; 28.6%) and most patients were male (n=6; 85.7%). Interestingly, IHC was negative for LTAg as well as VP1 in all patients. Therefore, it must also be stated that both virus proteins were negative in IHC for acute as well as chronic GvHD of the skin. Figure 1 illustrates our results in one patient with a Hematoxylin and Eosin (H&E) staining of the acute GvHD as well as the IHCs for LTAg and VP1. This patient was a male patient in his 6^th^ decade with an NHL as underlying disease. He had already BKPyV viruria with 3427 copies/mL at inpatients admission for alloSCT and had nearly stable BKPyV viruria with round about 3000 copies/mL during his inpatient treatment of 40 days. Additionally, he developed acute renal injury at day 20 after alloSCT (AKIN III, RIFLE injury). Biopsy proven acute GvHD of the skin was diagnosed histologically at day 99 after alloSCT, when BKPyV viruria was also present as mentioned above.

**Figure 1:**
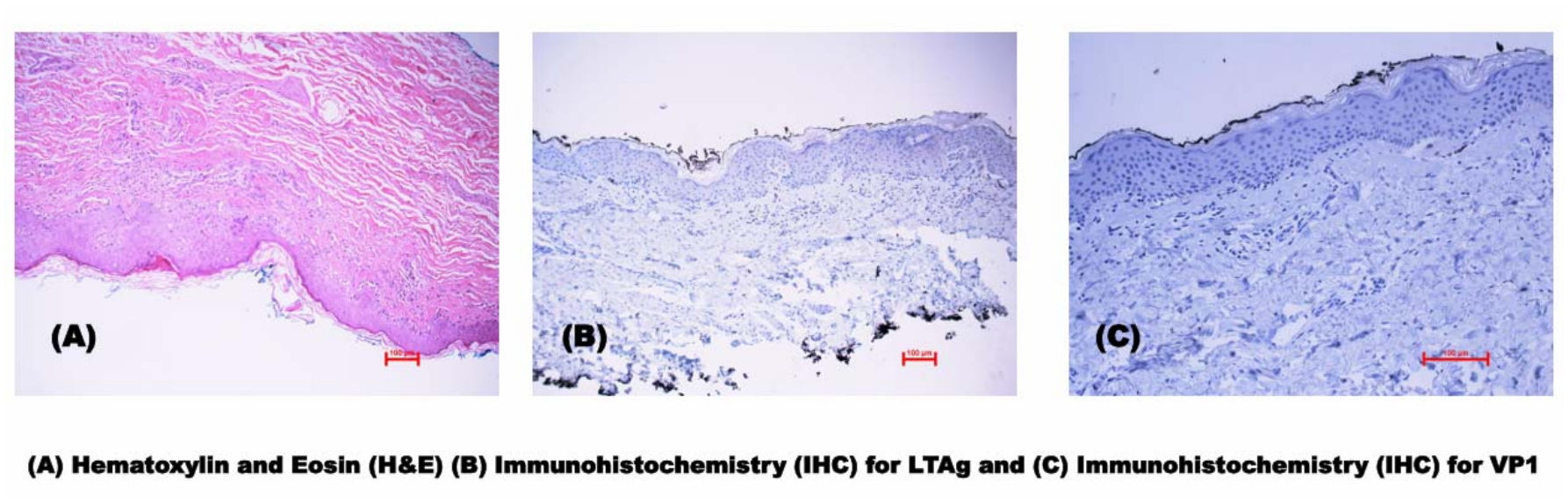
Male patient with NHL; staining of acute GvHD of the skin.

## Discussion

We conducted a pilot immunohistochemical study about the relationship of BKPyV and GvHD. Early and late virus proteins (LTAg and VP1) were not detectable immuno-histochemically in histological-proven GvHD of the skin. Therefore, an association between the virus reactivation/infection with GvHD is unlikely and further investigations for causality are not constructive. However, clearly stated it means that there is no association between skin GvHD and both virus proteins, but further research should focus on immunological factors and immunosuppression to be efficient, e. g. acute GvHD usually requires intensive immunosuppression and thereby increasing the risk of viral infections [13]. Furthermore, these investigations are clinically important since BKPyV-associated diseases, like HC, nephropathy or urothelial cancer, can lead to severe morbidity and even mortality in alloSCT [21; 22; 24].

Unfortunately, our study is not without several limitations. Firstly, it is only a pilot study with a very small sample size leading to selection bias. Secondly, we only used one method as well as no molecular detection methods to detect the virus in the GvHD tissue, like initially planned in our study protocol. Thirdly, we had only probes from GvHD of the skin available at the timepoints of BKPyV viruria, so our conclusion primarily applies for skin GvHD, which also was not consequently graded during all treatment phases. Additionally, due to the nature of a brief report and the fact we found no association and therefore decided not to perform further investigations, there is no detailed analysis for e. g. the timeline or detailed information of the GvHD included or viral loads. Strictly, we must state that there is only no association between the virus protein LTAg and VP1 and skin GvHD. Fourthly, in all patients BKPyV was not assessed prior to alloSCT. Lastly, we had no tissue available from the bladder or kidney from those patients. This is of special interest since the BKPyV persists in urothelium and tubulus cells. However, in this context other reasons than BKPyV nephropathy for renal impairment in patients with alloSCT also must be discussed, like CMV infec-tion/reactivation or nephrotoxic drugs, e. g. cyclosporine A [25; 26; 27; 28; 29].

Nevertheless and at least to our knowledge, this is the first investigation asking for an association between BKPyV virus reactivation/infection and GvHD. Despite the fact that this is relation was unlikely before, due to an obvious immunological explanation (GvHD is treated with immunosuppressive medication in first line) this study provides some more evidence, especially since the patients were identified from prospective studies. Additionally, also negative results are of high scientific value, because research than could be more focused and efficient, e. g. our working group also decided against further investigations.

In summary, there might be no evidence for an association between BKPyV reaction/infection and skin GvHD and so further studies should focus on immunological factors for the described association between the virus and GvHD, identification of risk factors for both diseases and the identification of a causal therapies for BKPyV infections and its associated diseases.

## Conclusions

There is no evidence for an association between the viral proteins LTAg and VP1 of BKPyV and skin GvHD in immunohistochemistry. Due to the pilot character of our investigation, the results should be interpreted with great caution. Multi-center studies could be useful for further investigations.

## Data Availability

All data produced in the present study are available upon reasonable request to the authors

## Statements

### Statement of ethics

#### Study approval statement

Before starting the study, we obtained the approval of the local ethics review board at the University Medical Center Greifswald (BB 264/25 from December 21, 2025).

#### Consent to participate statement

All patients gave written informed consent.

### Authors Contribution

Data collection: A. S., T. N., W. K., S. R. and L. S. (all authors); Conception and administration of the data: A. S., S.R. and L. S.; validation: A. S. and L. S., formal analysis: A. S. and L. S., writing and editing the manuscript: A. S. and L. S., critical review of the manuscript: A. S., T. N., W. K., S. R. and L. S. (all authors)

### Data availability statement

All data generated in this study are shown in this publication. Further enquiries can be directed to the corresponding author.

